# Integrating genetics with newborn metabolomics in infantile hypertrophic pyloric stenosis

**DOI:** 10.1101/2020.02.22.20026724

**Authors:** João Fadista, Line Skotte, Julie Courraud, Frank Geller, Sanne Gørtz, Jan Wohlfahrt, Mads Melbye, Arieh S. Cohen, Bjarke Feenstra

**Author notes:** Address correspondence to: João Fadista, PhD. Department of Epidemiology Research, Statens Serum Institut, Artillerivej 5, 2300 Copenhagen, Denmark. **Author Contributions:** JF designed the study, carried out the initial analyses and drafted the initial manuscript. JW, FG and SG reviewed and revised the manuscript and gave significant statistical contributions to the design and analyses plan. LS and JC designed the data collection instruments, collected data, carried out the initial analyses, and reviewed and revised the manuscript. MM and ASC conceptualized and designed the study, and critically reviewed the manuscript for important intellectual content. BF conceptualized, designed the study, coordinated and supervised data collection, and critically reviewed the manuscript for important intellectual content. All authors approved the final manuscript as submitted and agree to be accountable for all aspects of the work. **Financial Disclosure:** The authors have no financial relationships relevant to this article to disclose.

## Abstract

**Background:** Infantile hypertrophic pyloric stenosis (IHPS) is caused by hypertrophy of the pyloric sphincter smooth muscle. Building on a previously reported association between IHPS and lipid metabolism, we aimed to (1) investigate associations between IHPS and a wide array of lipid-centric targeted metabolites in newborns, and (2) address causality of the associations by integrating genetic data.

**Methods:** Dried blood spots were taken from 267 pairs of IHPS cases and controls matched by sex and day of birth. A mixed effects linear regression model was used to evaluate associations between Biocrates p400 Kit selected 148 metabolites and IHPS in a matched case-control design.

**Results:** Seven metabolites showed significantly lower levels in IHPS cases. Levels of the top associated metabolite, phosphatidylcholine PC(38:4), were significantly correlated with levels of the remaining six metabolites (P<2.30 ⨯ 10^−12^). Associations were driven by case-control pairs with older age at sampling. IHPS cases had more diagnoses for neonatal difficulty in feeding at breast (P = 6.15 ⨯ 10^−3^). Genetic variants explaining a large proportion of variance in the levels of PC(38:4) did not associate with IHPS.

**Conclusions:** We detected lower levels of certain metabolites in IHPS, possibly reflecting different feeding patterns in the first days of life.

**Key points:**

– Our previous GWAS of IHPS identified genetic associations between lipid metabolism and IHPS.
– Building on that finding, this study investigated associations between a wide array of lipid-centric metabolites in newborns and IHPS.
– Newborns who later developed IHPS had lower levels of certain metabolites. The associations were driven by individuals born at a later age at sampling, suggesting different feeding patterns after birth.

## INTRODUCTION

Infantile hypertrophic pyloric stenosis (IHPS), a disease typically appearing between two and eight weeks after birth, is characterized by hypertrophy of the pyloric sphincter smooth muscle, which leads to obstruction of the gastric outlet^1^. IHPS has a population incidence between 1 and 4 per 1000 live births^2,3^, a 4-to 5-fold higher risk in boys than girls^1^, and is the most common disease requiring surgery in the first months of life^4^. Despite its well-established clinical presentation, diagnosis and treatment, IHPS etiology remains incompletely understood, with both genetic and environmental factors contributing to disease pathogenesis. IHPS familial aggregation of 20-fold increased risk among siblings^5^, twin heritability estimates of 87%^5^ and genome wide SNP heritability of 30%^6^ suggest a strong genetic component for IHPS. Furthermore, perinatal risk factors such as exposure to macrolide antibiotics^7^, being first-born, preterm, delivery by cesarean section^8^, and bottle-feeding^9,10^ highlight the importance of modifiable early environmental exposures on the risk of IHPS^11^.

Previous findings from our group and others showed an association between IHPS and lipid metabolism. Briefly, we detected genetic variants at the *APOA1* locus in which known cholesterol-lowering alleles were associated with increased risk of IHPS^12^. Moreover, in the same study we found lower levels of total cholesterol in umbilical cord blood in cases vs. controls^12^. In our most recent genome-wide association study (GWAS) meta-analysis of IHPS, we also detected a genome-wide positive genetic correlation with high-density lipoprotein (HDL) cholesterol and a negative correlation with very low-density lipoprotein cholesterol (VLDL)^6^. Moreover, 14% of children with Smith–Lemli– Opitz syndrome (SLOS), a malformation syndrome caused by an inborn defect of cholesterol biosynthesis^13^, are reported to have IHPS^14^.

Following on these previous findings, this study had two goals. First, we wanted to investigate associations between IHPS and lipid-centric targeted metabolites in the blood of neonates from dried blood spot samples obtained through the routine Danish neonatal screening program^15^. Second, we wanted to address the question of whether detected differences in metabolite levels were likely to be driven by genetic differences between IHPS cases and controls or by differences in early life feeding patterns.

## METHODS

### Study Participants

Study participants belong to a Danish ancestry cohort previously described in our genome-wide meta-analysis of IHPS^6^. Briefly, IHPS cases were defined as having a pyloromyotomy in their first year of life, were singletons born in Denmark with parents and grandparents born in Northwestern Europe, and were born with no severe pregnancy complications nor major congenital malformations. For controls we used the same selection criteria as for cases, but without any IHPS diagnosis or surgery code. Our initial sample size comprised a total of 588 samples taken from dried blood spots (DBS) (294 pairs of cases and controls matched by sex and day of birth), of which we removed sequentially the following samples: (a) 2 pairs due to low sample quality in the cases; (b) 19 pairs due to at least one of the samples in each pair not passing standard GWAS QC (e.g. genetic variant missingness, ancestry outlier or genetic relatedness)^6^; (c) 5 pairs in which the controls had missing parity information; and (d) 1 pair in which the case had missing gestational age information. After this filtering, the discovery cohort comprised 267 pairs of IHPS cases and controls matched by sex and day of birth (range from 1997 to 2014) (Table 1, Supplemental Figure 1). The 267 IHPS cases represent a random 1/3 of all surgery confirmed cases in Denmark in the first year of life during the study period of 1997 up to 2014 (http://biobanks.dk/). All 534 samples were selected from DBS taken a few days after birth and stored in the Danish Neonatal Screening Biobank^15^. Sex, date of birth, gestational age, maternal age, Apgar scores and parity information were extracted from the Danish Medical Birth Register^17^, while diagnosis and surgery codes were extracted from the Danish National Patient Register^18^. Information on DBS sampling days after birth was available since 2006 in the Danish Neonatal Screening database. Sample summary characteristics are described in Table 1. This study was approved by the Danish Scientific Ethics Committee and the Danish Data Protection Agency. An exemption from obtaining informed consent from participants was given as this research project was based on samples from biobank material (H-4-2013-055).

**Table 1.** Study Sample Characteristics.

|  | **Cases (N=267)** | **Controls (N=267)** |
| --- | --- | --- |
| **Boys, No. (%)** | 233 (87.3) | 233 (87.3) |
| **Year of birth (range)** | 1997 - 2014 | 1997 - 2014 |
| **Year of birth, mean (SD)** | 2008 (4.6) | 2008 (4.6) |
| **Maternal age, mean (SD), years** | 29.8 (5.3) | 30.5 (4.8) |
| **Gestational age, mean (SD), weeks** | 39.8 (1.7) | 40.0 (1.4) |
| **Parity, mean (SD)** | 1.7 (1.0) | 1.8 (0.8) |
| **Apgar scores, mean (SD)** | 9.8 (0.7) | 9.9 (0.4) |
| **Age at DBS sampling, mean (SD), days** | 3.3 (2.2) | 3.2 (1.8) |
| **Age at diagnosis, mean (SD), days** | 36.8 (18.1) |  |

**Figure 1.**
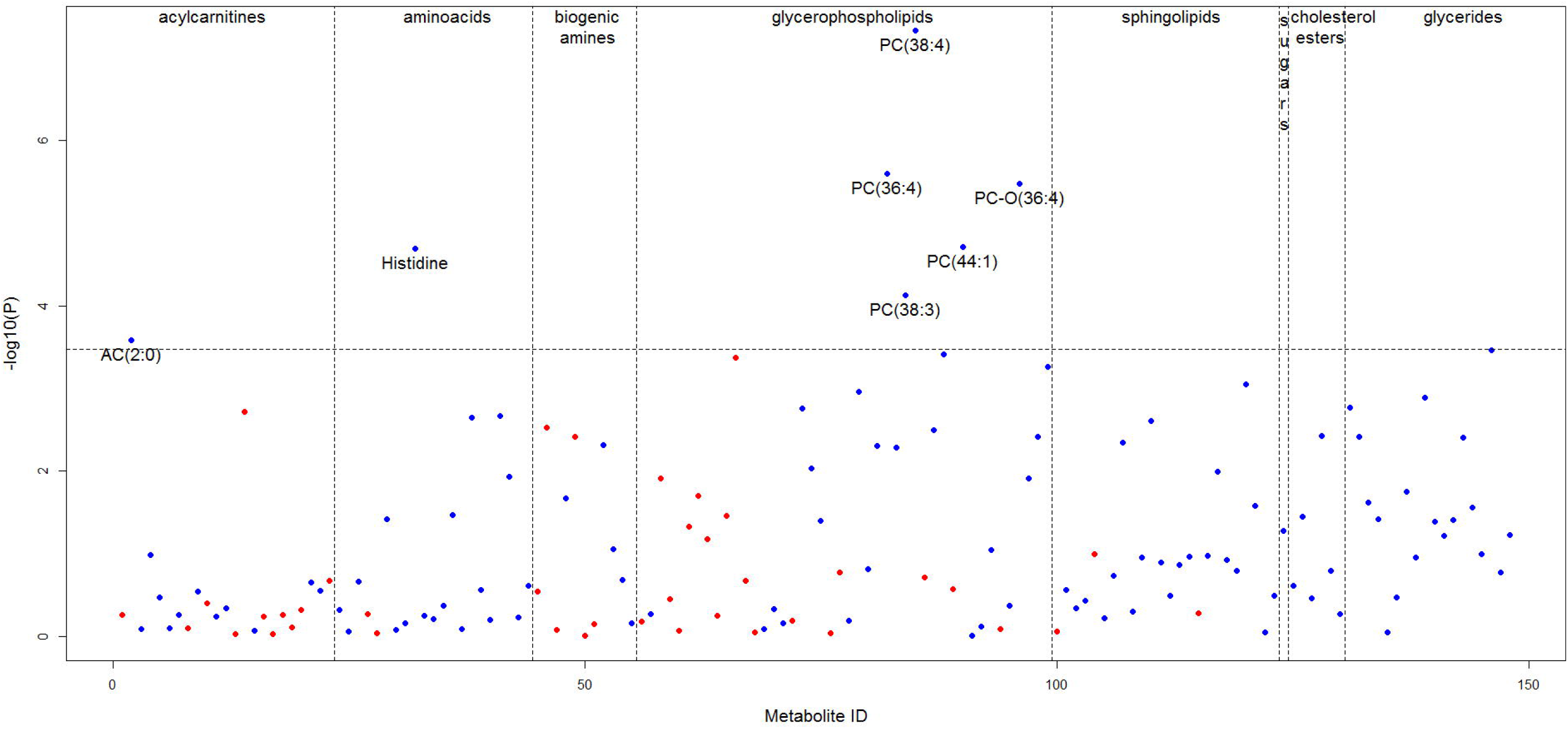
IHPS association with 148 metabolites tested. Dashed vertical lines separate different metabolite classes. The dashed horizontal line marks the Bonferroni-adjusted significance level of P=0.05/148. Blue and red dots represent metabolites with lower and higher levels in IHPS cases vs. controls, respectively. Case/Control normalized concentration fold-changes are between 0.67 and 1.26.

### Metabolite profiling and nomenclature

The DBS samples consist of whole blood blotted onto filter paper, dried at room temperature for at least 3 hours before being sent by mail for newborn screening at Statens Serum Institut. After newborn screening, the samples were then stored at −20°C in the Danish National Biobank. On the day of the analysis, a 3.2-mm punch was collected using a Panthera-PuncherTM 9 blood spot punching system (PerkinElmer, Waltham, MA, USA) directly into the 96-well kit plates.

Targeted metabolite quantification followed the manufacturer’s user manual for the AbsoluteIDQ® p400 Kit (Biocrates Life Sciences AG, Innsbruck, Austria), adapted for DBS analysis by the manufacturer^19^. The kit has a high-performance liquid chromatography (LC) separation step and a flow injection analysis (FIA) step, both followed by mass spectrometry (MS) analyses. Mass detection and compound identification are performed by multiple reaction monitoring. The method of the AbsoluteIDQ® p400 kit has comparable results to the AbsoluteIDQ® p180 kit^20^, a kit with reported high interlaboratory reproducibility^21^, and validated for human blood plasma and DBS^22^. The AbsoluteIDQ® p400 kit allows simultaneous quantification of 408 metabolites comprising 55 acylcarnitines, 21 amino acids, 21 biogenic amines, 196 glycerophospholipids (24 lysophosphatidylcholines and 172 phosphatidylcholines), 40 sphingolipids (31 sphingomyelins and 9 ceramides), hexoses (about 90-95 % glucose), 14 cholesterol esters and 60 glycerides (18 diglycerides and 42 triglycerides). Lipid metabolites are abbreviated as follows: acylcarnitines as AC(X:Y); hydroxylacylcarnitines as AC(X:Y-OH); dicarboxylacylcarnitines as AC(X:Y-DC); phosphatidylcholines as PC(X:Y); lysophosphatidylcholines as LPC(X:Y); sphingomyelins as SM(X:Y); ceramides as Cer(X:Y); cholesterol esters as CE(X:Y); diglycerides as DG(X:Y) and triglycerides as TG(X:Y). Each lipid metabolite is mentioned as X:Y, with X being the number of carbon atoms and Y the number of double bonds. Quality was controlled on each plate by the injection of three paper-based blanks and three quality control (QC) levels (QC1-3), of which the medium level (QC2) was injected in five replicates. Absolute concentrations were calculated based on 7-point calibration curves for LC-MS metabolites and 1-point calibration for FIA-MS metabolites.

The equipment (Thermo Fisher Scientific, Waltham, MA, USA) consisted of a Combi PAL HTS TMO autosampler, a LX-2 LC system with two injectors, each connected to an Ultimate 3000 Dionex RS pump and to the MS through a Transcend II Valve Interface Module. MS data was acquired on a QExactive with a heated electrospray ionization source. All solvents that were not provided with the kit were Optima(tm) LC/MS grade and purchased from Thermo Fisher Scientific. The instruments were controlled using TraceFinder 4.1 Clinical Research, Aria MX V2.2 and QExactive tune software V2.8 SP1.

We first injected extracts for LC-MS measurements on one dedicated injector (amino acids, biogenic amines). FIA was conducted the next day on the second dedicated injector (acylcarnitines, glycerophospholipids, sphingolipids, hexoses, cholesterol esters, glycerides). Eight 96-well plates were run, with wells measured in a fixed order (from #1 to #96). A matched case and control would always be on the same plate but not necessarily injected sequentially. LC-MS data was pre-processed using Xcalibur 4.1, and all data integrated in Biocrates MetIDQ Carbon-2793 software for quality assessment, quantification of metabolite concentrations by reference to appropriate internal standards, and to do inter- and intra-plate normalization using the median value of the five QC2 replicated injections of each plate in comparison with their target values (see below).

### Statistical Analysis

For each metabolite m and plate p, the limit-of-detection (LOD_m,p_) was defined as the median non-normalized values of metabolite m for paper-based blanks (done in triplicate for each plate) on the relevant plate p. This means that LODs were calculated separately for each plate. We excluded a metabolite m, if more than 20% of non-normalized concentrations in cases and controls were ≤ LOD. MetIDQs data normalization procedure was implemented in R^23^, to scale out differences between plates. Briefly, the median value per metabolite per plate of the five replicated injections of the QC2 (A_m,p_) was calculated (only for cells above LOD) and divided that by the metabolite target value (TV_m_) of each metabolite, available in the MetIDQ database, leading to the correction factor by plate (A_m,p_/TV_m_ = C_m,p_). Then to normalize the data, the raw concentration values of all samples were divided by the plate-specific correction factor: Norm_m,p_ = Raw_m,p_/C_m,p_. We also did metabolite filtering by coefficient of variation (CV) based on the normalized values of the QC2 replicates. For each metabolite m, we calculated its CV for all plates together, i.e. for 5 QC2 samples and 8 plates, the CV was based on 40 observations. If CV_m_ > 25% we excluded metabolite m. Thus, of the 408 metabolites measured with the kit, we therefore selected 148 metabolites that were above the limit of detection (LOD) in at least 80% of the samples (in cases or controls separately) and had a coefficient of variation of the QC2 replicates below 25%.

Due to the non-normal distribution of metabolite concentrations (reported in µM), we quantile transformed the normalized metabolite concentrations to a standard normal distribution prior to all statistical analyses. Estimation of association of metabolite levels with IHPS was performed with a linear mixed-effects model, implemented in the lmer function from the lme4 R package^24^, adjusted for sex (as factor), year of birth (as factor), parity, and gestational age (in weeks) as fixed effects, and IHPS case-control pair as a random effect. A statistically significant association was defined as P < 0.05/148, to adjust for the 148 metabolites tested (Bonferroni correction).

### Diagnosis codes and co-occurrence with IHPS

All International Classification of Diseases (ICD) diagnosis codes from our 267 pairs of IHPS cases and controls were extracted from the Danish National Patient Register^18^. Fisher’s exact test for count data implemented in R^23^ was used to calculate the odds-ratio of occurrence of the ICD-code P92.5 in IHPS cases vs. controls.

### Genetic variants associated with metabolite levels

Assessing the effect of *FADS1* genetic variant rs174547^25-27^ on IHPS was done doing a lookup in our previous genome-wide meta-analysis of IHPS^6^. The association of rs174547 with metabolite levels in our cohort was determined in a linear model adjusted for IHPS status, sex, year of birth (as factor), parity, and gestational age (in weeks). Stratified analysis was also conducted separately for individuals born from 2009 onwards (lower age at sampling) and for individuals born before 2009 (higher age at sampling).

## RESULTS

### Description of the cohort

A cohort comprising 267 pairs of IHPS cases and controls matched by sex and day of birth was sampled from DBS samples taken a few days after birth (Methods, Supplementary Methods). Key characteristics of the cohort are described in Table 1 and Methods.

### Targeted metabolomics and IHPS association

Based on our previous findings, in which we detected an association between IHPS and lipid metabolism^6,12^, we chose a lipid-centric targeted quantitative metabolomics platform for metabolic profiling (Methods, Supplementary Methods). Of the 408 metabolites measured with the Biocrates p400 Kit, 148 metabolites passed our quality curation (Methods, Supplementary Methods). Using a mixed effects linear regression model (Methods), seven of the 148 selected metabolites showed significantly lower levels in IHPS cases (Table 2, Figure 1, Supplemental Table 1). The seven metabolites included acylcarnitine AC(2:0), the amino acid histidine and five phosphatidylcholines (PC(38:4), PC(36:4), PC-O(36:4), PC(44:1) and PC(38:3)) (Table 2, Figure 1). The top IHPS associated metabolite was phosphatidylcholine PC(38:4) (P= 4.68 × 10^−8^) (Figure 2), while the concentrations of the remaining six metabolites showed significant positive correlation with PC(38:4) (all correlations with P< 2.30 × 10^−12^; test for correlation between paired samples using Pearson’s product moment correlation coefficient) (Figure 3). Since the year 2009 marked a change of policy of the Danish neonatal screening program regarding age at sampling of DBS samples, from a median of 6 days (5-8 days; 1^st^-3^rd^ quartiles) to 2 days (2-3 days; 1^st^-3^rd^ quartiles) after birth (Supplemental Figure 2), we decided to also test the association of metabolite levels with IHPS for individuals born before 2009 and from 2009 onwards, separately. After correction for multiple comparisons, significant association of metabolite levels with IHPS were only detected for individuals born before 2009 but not from 2009 onwards (Table 2, Figure 2, Supplemental Tables 2-3). We found no evidence of association between sex and levels of PC(38:4) (sex effect in full model described above: P=0.59) (Supplemental Figure 3), and levels of the other six IHPS associated metabolites (results not shown).

**Table 2.** Associations of metabolites with IHPS. Results are shown for all pairs and stratified into pairs with year of birth <2009 or ≥2009. Metabolites are ordered by *P* value in the main analysis including all birth years.. The effect size Beta is with respect to the quantile transformed normalized concentration levels of each metabolite (Methods). *P* values of the metabolite associations significant after Bonferroni correction are highlighted in bold. Metabolite nomenclature is available in Methods section. YOB, Year of birth. SE, standard error. P, P value of the association. d, days.

| Metabolites | 267 IHPS case/control pairs |  |  | 98 IHPS case/control pairs |  |  | 169 IHPS case/control pairs |  |  |
| --- | --- | --- | --- | --- | --- | --- | --- | --- | --- |
|  | (All YOB) |  |  | (YOB<2009; median age=6d) |  |  | (YOB≥2009; median age=2d) |  |  |
|  | Beta | SE | P | Beta | SE | P | Beta | SE | P |
| PC(38:4) | -0.42 | 0.07 | **4.68E-08** | -0.71 | 0.12 | **1.28E-08** | -0.25 | 0.09 | 9.35E-03 |
| PC(36:4) | -0.36 | 0.07 | **2.51E-06** | -0.59 | 0.13 | **1.46E-05** | -0.23 | 0.09 | 1.51E-02 |
| PC-O(36:4) | -0.33 | 0.07 | **3.29E-06** | -0.68 | 0.12 | **3.44E-07** | -0.13 | 0.08 | 1.10E-01 |
| PC(44:1) | -0.33 | 0.08 | **1.94E-05** | -0.44 | 0.13 | 1.08E-03 | -0.28 | 0.10 | 3.95E-03 |
| Histidine | -0.34 | 0.08 | **2.01E-05** | -0.57 | 0.14 | **9.25E-05** | -0.22 | 0.09 | 2.17E-02 |
| PC(38:3) | -0.32 | 0.08 | **7.39E-05** | -0.59 | 0.12 | **5.31E-06** | -0.16 | 0.10 | 1.34E-01 |
| AC(2:0) | -0.25 | 0.07 | **2.60E-04** | -0.39 | 0.10 | **1.49E-04** | -0.17 | 0.09 | 6.42E-02 |

**Figure 2.**
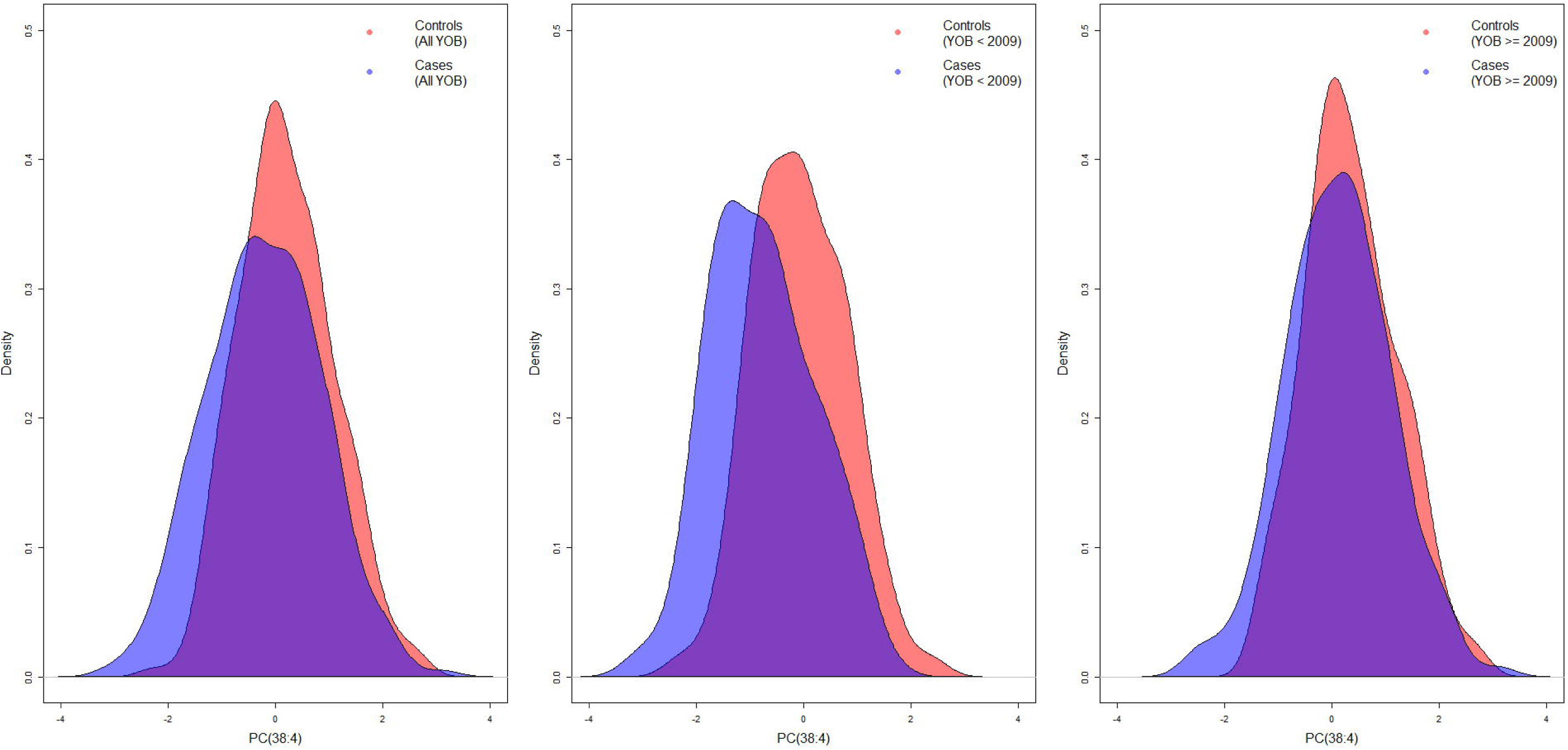
Density plot of the distribution of the quantile transformed normalized concentration of PC(38:4) in our cohort of 267 pairs of IHPS cases and controls.

**Figure 3.**
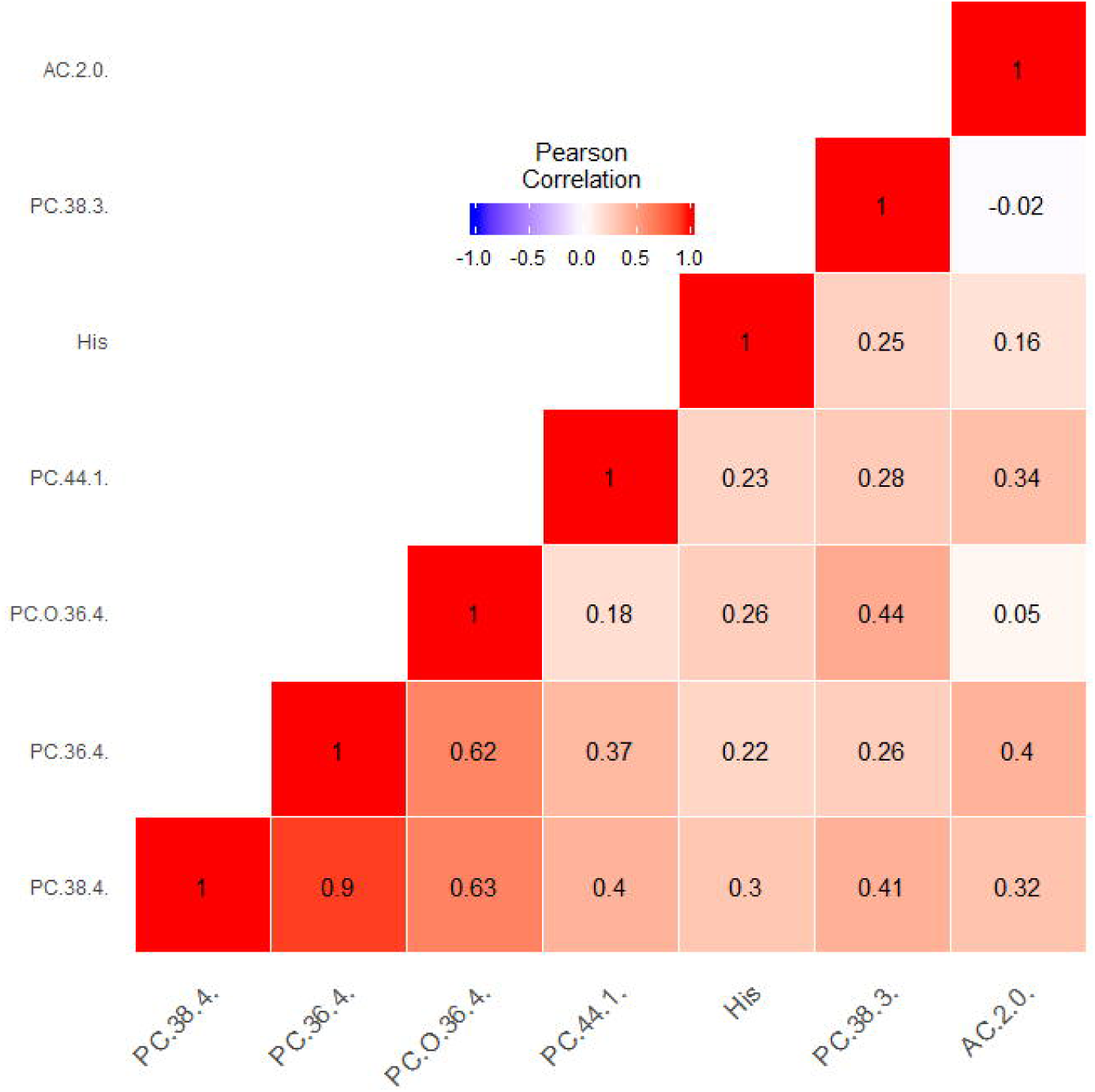
Pairwise complete Pearson correlation between the quantile transformed normalized concentrations of the seven significantly IHPS associated metabolites.

### IHPS-associated metabolites may tag different feeding patterns between cases and controls

Different lines of evidence indicate that the concentration of the IHPS-associated metabolite PC(38:4) may tag different feeding patterns between cases and controls in the first days of life. Serum levels of these metabolites are influenced by food intake and genetics. In adults, 11% of the variance in PC(38:4) levels is explained by genetic variation in *FADS1* gene^27^. The *FADS1* gene codes for the fatty acid delta-5 desaturase, a key enzyme in the metabolism of long-chain polyunsaturated omega-3 and omega-6 fatty acids, which catalyzes desaturation of PC(38:3) to PC(38:4)^25-27^. In our neonatal cohort, genetic variation in *FADS1* gene was associated with the ratio of PC(38:4)/PC(38:3) (rs174547 P= 5.83 ⨯ 10^−10^), with PC(38:3) (rs174547 P= 1.05 ⨯ 10^−5^) but not with PC(38:4) (rs174547 P= 0.30) (Supplemental Figure 4). For individuals born from 2009 onwards (lower age at sampling), genetic variation in *FADS1* gene was also associated with the ratio of PC(38:4)/PC(38:3) (rs174547 P= 1.27 ⨯ 10^−11^), with PC(38:3) (rs174547 P= 2.92 ⨯ 10^−6^) and also not with PC(38:4) (rs174547 P= 0.30). For individuals born before 2009 (higher age at sampling), genetic variation in *FADS1* was not associated with the ratio of PC(38:4)/PC(38:3) (rs174547 P= 0.17), PC(38:3) (rs174547 P= 0.46) nor with PC(38:4) (rs174547 P= 0.61). To test if the genetically determined levels of PC(38:4) have a causal role on IHPS etiology, we tested the influence of the *FADS1* gene cluster polymorphisms on IHPS risk. In our GWAS data^6^, genetic variation in *FADS1* was not associated with IHPS (sentinel SNP rs174547 P=0.90, Supplemental Figure 5). This suggests that other factors influencing PC(38:4) levels underlie the IHPS association, and feeding pattern could be one such factor. Although we do not have general information on feeding patterns in our cohort, a larger number of IHPS cases than controls had a diagnosis code for neonatal difficulty in feeding at breast (ICD-10 code P92.5) in the Danish National Patient Register (odds ratio [OR], 4.46; 95% confidence interval [CI]: 1.43-18.46; P = 6.15 × 10^−3^) (Supplemental Table 4, Methods).

## DISCUSSION

In this case-control metabolomics study of IHPS, we identified seven metabolites with significantly lower levels in newborns who later developed the disease. The metabolites included five PCs, acylcarnitine AC(2:0), and the amino acid histidine. The associations were driven by 98 case-control pairs born before 2009, when median age at sampling was 6 days. No significant associations were seen in the 169 case-control pairs born in 2009 or later, when median age at sampling was 2 days.

Genetic variation known to affect levels of our sentinel IHPS significant metabolite PC(38:4) in blood of adults^27^ was not associated in our neonatal cohort, nor was associated with IHPS in our previous GWAS meta-analysis^6^. This suggests non-causality of the genetically determined levels of the metabolite PC(38:4) with IHPS, requiring another explanation for the observed association between PC(38:4) levels and IHPS. The fact that IHPS cases had more diagnoses for neonatal difficulty in feeding at breast than controls supports the hypothesis that a higher proportion of cases were primarily bottle-fed. Furthermore, the phosphatidylcholine PC(38:4) is known to be at higher levels in plasma^28^ and dried blood spot^29^ samples from breast-fed infants compared to bottle-fed infants, and bottle-feeding is a well-known risk factor for IHPS^9-11^. Thus, IHPS risk could be exacerbated by introducing bottle-feeding, which may induce lower levels of phosphatidylcholines, essential in cholesterol metabolism^30^. This is in line with the argument that, right after birth, newborn’s milk intake (whether formula or breast-milk) is very low but increases rapidly within the first week of life^31,32^, which could explain the significant metabolite associations only for individuals born before 2009, as they were sampled at a later age. Moreover, the fact that the ratio of PC(38:4)/PC(38:3) was significantly associated with *FADS1* genetic variation for individuals born from 2009 (samples about 2 days after birth) but not for individuals born before 2009 (sampled at a later age) suggests that the effects of feeding patterns about one week after birth influence the ratio so much that the effect of *FADS1* genetic variation is masked. The association between PC(38:4) levels and IHPS (seen about 6 days after birth) is therefore more likely to be attributable to feeding differences between IHPS cases and controls.

Similar to phosphatidylcholines, histidine and the short-chain acylcarnitine AC(2:0) were also detected to be at significant lower concentrations in IHPS (Figure 1, Table 2, Supplemental Figures 6-7). The lower histidine concentration levels in IHPS cases could be further evidence that feeding differences exist between cases and controls. Histidine being an essential amino acid^33^, it cannot be synthesized *de novo* in humans and therefore, needs to be supplied in the diet. Moreover, histidine content is known to be higher in breast milk compared to standard liquid infant formulas^34^ (preferred formulas in the first days of life), suggesting that controls had a higher degree of breast-feeding than cases. Regarding AC(2:0), it has been reported that infants who are breast-fed have higher concentrations of this metabolite in their plasma compared to bottle-fed infants^35^, which further suggests the hypothesis that controls are being more breast-fed compared to cases.

Despite the epidemiological evidence that boys have a 4-to 5-fold higher IHPS risk than girls^1^, and that sex differences exist in serum lipids and lipoproteins at birth^36^, we did not detect sex differences in the concentrations of the IHPS associated metabolites.

Since our results seem to reflect mainly the influence of feeding practice on IHPS risk, our previous findings of lower levels of total cholesterol (TC) in umbilical cord blood in cases vs. controls^12^ and positive genetic correlation of IHPS with HDL cholesterol^6^ and negative genetic correlation with VLDL^6^ could not be explained by the results of the current study. However, together with the reported high incidence of IHPS in children with Smith–Lemli–Opitz syndrome (SLOS)^14^, caused by a cholesterol biosynthesis defect^13^, it appears that both genetic and environmental factors can contribute to the pathophysiology of IHPS through disturbances of the lipid metabolism of newborns.

Our study had some limitations, including the use of small amounts of sample material from DBS samples and the use of a targeted approach in which other relevant metabolites could have been overlooked. We also did not have general information on breast-feeding vs. formula feeding so we were not able to study the feeding effect directly. In addition, genetic variation known to associate with levels of metabolites queried here is only known in adults, and so we might have missed genetic associations specific to newborn metabolite levels. Strengths of the study include the use of a well-established targeted metabolomics platform (Biocrates) to investigate the association between IHPS and 148 metabolites from DBS samples taken a few days after birth. To our knowledge, this is the first time that extensive newborn metabolomics profiling is being used to study IHPS. Another strength of the study is the use of well-defined codes from nation-wide Danish registers to define outcome, exclusions, and covariates. Finally, our study design in which we performed 1-to-1 matching of IHPS cases and controls based on sex and exact day of birth is also a clear strength of the study.

Our observations have implications for ongoing studies of complex diseases in which both genetics and environmental factors play a role in disease etiology. In time, a detailed molecular dissection of IHPS risk factors could potentially be used as a basis for more rapid identification of particularly susceptible newborns. While our study represents a step in that direction, further efforts are needed to replicate our findings in other populations and to better understand the interplay of genetics and early feeding patterns and consequent progression to IHPS. Specifically, the use of wide lipid profiling in nutritional intervention studies could help to unravel causal relationships and underlying mechanisms. Moreover, since AC(2:0) is already present on the list of most of newborn screening panels worldwide^37^, future work could address the predictive power of AC(2:0) as a newborn screening test for IHPS.

## CONCLUSIONS

Although IHPS is highly heritable^5,6^, it is a complex disease that does not have Mendelian transmission through families. Common genetic variants^6,12,16^ and environmental risk factors^7-11^ have been previously discovered, but IHPS etiology remains incompletely understood. In this case-control metabolomics study of IHPS we have detected lower levels of PC(38:4), together with correlated PCs, histidine and AC(2:0), in IHPS. Several lines of evidence suggest that environmental factors affecting these metabolites such as feeding pattern in the first days of life may underlie the associations. Further understanding of the complex interplay of genetics and early life environmental factors is critical to the understanding of the pathophysiology, diagnosis and possible treatment of IHPS, and can serve as a model for other complex diseases.

## Data Availability

All data is available upon request.

## Acknowledgments

We would like to thank Susan Svane Laursen for running the metabolomics analyses and for curating the metadata necessary for the study.

## Author Contributions

JF designed the study, carried out the initial analyses and drafted the initial manuscript. JW, FG and SG reviewed and revised the manuscript and gave significant statistical contributions to the design and analyses plan. LS and JC designed the data collection instruments, collected data, carried out the initial analyses, and reviewed and revised the manuscript. MM and ASC conceptualized and designed the study, and critically reviewed the manuscript for important intellectual content. BF conceptualized, designed the study, coordinated and supervised data collection, and critically reviewed the manuscript for important intellectual content. All authors approved the final manuscript as submitted and agree to be accountable for all aspects of the work.

## SUPPLEMENTARY TABLE LEGENDS

**Supplemental Table 1**. Main IHPS association results for the 148 metabolites tested.

**Supplemental Table 2**. Main IHPS association results for the 148 metabolites tested for individuals born before 2009.

**Supplemental Table 3**. Main IHPS association results for the 148 metabolites tested for individuals born in 2009 or later.

**Supplemental Table 4**. Number of IHPS cases and controls with and without the ICD-10 code P92.5 describing neonatal difficulty in feeding at breast.

## SUPPLEMENTARY FIGURE LEGENDS

**Supplemental Figure 1**. Participant flowchart.

**Supplemental Figure 2**. Boxplots of the distribution of age at sampling (in days) of dried blood spots, stratified by IHPS case/control status and year of birth (YOB) before 2009 and from 2009.

**Supplemental Figure 3**. Boxplots of the distribution of the quantile transformed normalized concentrations of PC(38:4), stratified by sex and IHPS case/control status.

**Supplemental Figure 4**. Boxplots showing the relationship between the genotype of SNP rs174547 and quantile transformed normalized concentrations of PC(38:4), PC(38:3) and the ratio PC(38:4)/PC(38:3).

**Supplemental Figure 5**. Regional association plot of the IHPS association results at the *FADS1* locus. Color-coded linkage disequilibrium is shown for the sentinel SNP rs174547. Linkage disequilibrium determined for the 1000 Genomes EUR population, November 2014 freeze, as described at http://locuszoom.sph.umich.edu/. The x-axis represents the genomic region (hg19 assembly) surrounding 400kb of rs174547, while the y-axis represents the strength of the association in −log10(P-value).

**Supplemental Figure 6**. Density plot of the distribution of the quantile transformed normalized concentration of Histidine in our cohort of 267 pairs of IHPS cases and controls.

**Supplemental Figure 7**. Density plot of the distribution of the quantile transformed normalized concentration of AC(2:0) in our cohort of 267 pairs of IHPS cases and controls.

